# Association of maternal and infant inflammation with neurodevelopment in HIV-exposed uninfected children in a South African birth cohort

**DOI:** 10.1101/2020.05.03.20089383

**Authors:** Tatum Sevenoaks, Catherine J. Wedderburn, Kirsten A. Donald, Whitney Barnett, Heather J. Zar, Dan J. Stein, Petrus J.W. Naudé

## Abstract

HIV-exposed uninfected (HEU) children may have altered immune regulation and poorer neurodevelopment outcomes compared to their HIV-unexposed (HU) counterparts. However, studies investigating the association of maternal and infant inflammation with neurodevelopment in HEU children are limited and longitudinal data are lacking. This study investigated serum inflammatory markers in HIV-infected *vs*. uninfected women during pregnancy and in their children, as well as associations with neurodevelopmental outcomes at two years of age in an African birth cohort study. A sub-group of mother-child dyads from the Drakenstein Child Health Study had serum inflammatory markers measured at ≈26 week’s gestation (n=77 HIV-infected mothers; n=190 HIV-uninfected mothers), at 6-10 weeks (n=63 HEU infants and n=159 HU infants) and at 24-28 months (n=77 HEU children and n=190 HU children). Serum inflammatory markers [granulocyte-macrophage colony-stimulating factor (GM-CSF), interferon-γ (IFN-γ), interleukin IL-1β, IL-2, IL-4, IL-5, IL-6, IL-7, IL-8, IL-10, IL-12p70, IL-13, tumor necrosis factor-*α* (TNF-*α*), neutrophil gelatinase-associated lipocalin (NGAL) and metalloproteinase-9 (MMP-9)] were analyzed with a multiplex bead array and ELISA assays. The Bayley Scales of Infant and Toddler Development, third edition, was used to assess neurodevelopment at 24-28 months. After correcting for multiple comparisons, HIV-infection during pregnancy was associated with lower serum levels of inflammatory markers in mothers at 26 weeks gestation (GM-CSF and MMP9, *p<0.05*) and HEU children at 6-10 weeks (IFN-γ and IL-1β, *p<0.01*), and at 24-28 months (IFN-γ, IL-1β, IL-2 and IL-4, *p<0.05*) compared to HIV-uninfected mothers and HU children. In HEU infants at 6-10 weeks, inflammatory markers (GM-CSF, IFN-γ, IL-10, IL-12p70, IL-1β, IL-2, IL-4, IL-6 and NGAL, *all p<0.05*) were associated with poorer motor function at two years of age. This is the first study to evaluate the associations of follow-up immune markers in HEU children with neurodevelopment. These findings suggest that maternal HIV infection is associated with immune dysregulation in mothers and their children through two years of age. An altered immune system in HEU infants is associated with poorer follow-up motor neurodevelopment. These data highlight the important role of the immune system in early neurodevelopment and provide a foundation for future research.

## 1. INTRODUCTION

Maternal Human Immunodeficiency Virus (HIV) infection affects maternal physiology and may have far-reaching consequences for the development of the exposed foetus and child. With wider access to antiretroviral therapy (ART) to prevent transmission of HIV from mother to child, the number of HIV-exposed uninfected (HEU) children is increasing. This is particularly relevant in South Africa, where the mother-to-child transmission rate dropped to <5% in 2018, resulting in a current HEU child population of 3 500 000 (UNAIDS, 2019). However, exposure to HIV *in utero* may impair the health of these children, including their neurodevelopment (Evans et al., 2016; McHenry et al., 2018; Wedderburn et al., 2019).

Several mechanisms have been proposed to explain the adverse outcomes in HEU children including; maternal and child immune dysregulation, ART toxicity, higher exposure to infectious diseases and socio-environmental factors. The impact of HIV infection on the adaptive immune system has been well described in adults (Mohan et al., 2014). However, few studies have examined maternal immunity in pregnancy in the context of HIV infection. Previous reports suggest that HIV impacts the maternal immune system by creating a pro-inflammatory environment, which is characterised predominantly by increased levels of certain cytokines, such as interleukin (IL-1, IL-6) and tumour necrosis factor-α (TNF-α) (Faye et al., 2007; Richardson and Weinberg, 2011; Sachdeva et al., 2008). Findings from studies have shown suppression of cytokine levels in HIV-infected pregnant women (Maharaj et al., 2017; Moussa et al., 2001). Further, altered immune regulation may be present in HEU children (Abu-Raya et al., 2016a). The vulnerability of the immune system in HEU children is also suggested by the increase in morbidity and mortality in HEU infants (Afran et al., 2014). However, data on HEU children inflammatory profiles are inconsistent, with evidence for both increased levels (Dirajlal-Fargo et al., 2019; Miyamoto et al., 2017; Prendergast et al., 2017) as well as reduced levels of cytokines in HEU children (Borges-Almeida et al., 2011; Chougnet et al., 2000).

A recent meta-analysis found poorer cognitive and motor neurodevelopment in HEU children (McHenry et al., 2018). Additionally, we have found adverse language outcomes at 2 years in HEU infants in the Drakenstein Child Health Study (DCHS) in South Africa (Wedderburn et al., 2019) but the biological mechanisms underlying this remain largely unknown. Studies suggest that maternal immune activation and increased levels of certain inflammatory markers during pregnancy may contribute to poor foetal brain development (Bilbo and Schwarz, 2012; Hsiao and Patterson, 2012; Morelli S, 2015; White et al., 2020). Due to the expanding population of HEU children and concerns about neurodevelopmental vulnerability in these children, the association between HIV, the immune system and neurodevelopment is a critical area of investigation.

The aims of this study were to investigate the impact of HIV on inflammatory profiles of pregnant women and their uninfected children through two years, and to assess whether these are associated with neurodevelopmental outcomes in the children at 2 years of age in the DCHS.

## 2. METHODS

### 2.1. Study participants

A sub-sample of randomly selected mother-child pairs (n=267) from the Drakenstein Child Health Study (DCHS), a large population-based birth cohort, as previously described, were included in this study (Stein et al., 2015; Zar et al., 2015). In brief, enrolment took place between 2012 and 2015 at two primary health care clinics, TC Newman and Mbekweni in Paarl, South Africa. All births occurred at Paarl Hospital. Pregnant women were enrolled if they were at least 18 years of age, planned to live in the region for at least 1 year and provided written informed consent (Stein et al., 2015; Zar et al., 2015).

This sub-study and the DCHS were approved by the Faculty of Health Sciences, Human Research Ethics Committee (HREC) of the University of Cape Town (HREC 401/2009 and HREC 648/2018).

### 2.2. Study procedures

#### 2.2.1 Variable measures and collection

Routine HIV testing of pregnant women was conducted to confirm HIV status in accordance with the Western Cape prevention of mother-to-child transmission (PMTCT) HIV guidelines (Pellowski et al., 2019). All HIV-exposed uninfected (HEU) children received HIV testing as per local guidelines, and all HEU children in the sub-sample were confirmed negative. Detection of HIV was done at 6 weeks by PCR and at 9 months and 18 months by rapid antibody, PCR or ELISA (Zar et al., 2015).

Maternal CD4 cell count and viral load during pregnancy were measured, with the result closest to 26 weeks’ gestation taken to coincide with the immune variables. Viral load was categorized as below the detectable limit with <40 copies/ml, detectable with 40-1000 copies/ml and unsuppressed with >1000 copies/ml. All HIV-infected mothers received antiretroviral therapy (ART) during pregnancy according to PMTCT guidelines at the time. All HEU infants received prophylaxis (nevirapine alone or combined with zidovudine) from birth (Pellowski et al., 2019).

Blood serum samples were taken at 26 weeks gestation for mothers and at 6-10 weeks and 24-28 months for children as outlined in the DCHS (Zar et al., 2015).

Sociodemographic information was collected using an interviewer administered questionnaire adapted from items used in the South African Stress and Health Study. Mothers self-reported employment status, education level, asset ownership, household income and clinic during an antenatal study visit between 28- and 32-weeks gestation (Stein et al., 2015; Zar et al., 2015).

Upon delivery detailed birth data was obtained, including mode of delivery, gestational age, infant sex, head circumference, infant length and infant birth weight. Prematurity was defined at less than 37 weeks gestation (Budree et al., 2017b). Maternal Body Mass Index (BMI) was determined at 6 weeks postpartum (Budree et al., 2017b). Self-reported information on child feeding practices was obtained at infant follow-up visits at 6-10 weeks, and 24-28 months of age. At 6-10 weeks infants were categorized as exclusively breastfed if mothers were still breastfeeding but neither solids nor formula had been introduced (Budree et al., 2017a; Wedderburn et al., 2019; Zar et al., 2015).

Maternal alcohol use during pregnancy was assessed using the Alcohol, Smoking, and Substance Involvement Screening Test; mothers were classified with either moderate-severe alcohol exposure *vs*. un-exposed (Stein et al., 2015). Infants and children were classified as being alcohol-exposed *in utero* if their mother was classified with moderate-severe alcohol exposure. Smoke exposure was measured at 26 weeks gestation using urine cotinine levels that were determined using the IMMULITE 1000 nicotine metabolite kit (Vanker et al., 2016). Mothers were considered a non-smoker if cotinine levels were <10ng/ml, a passive smoker with levels between 10-500ng/ml and an active smoker with levels >500ng/ml.

#### 2.2.2 Neurodevelopmental Assessment

Neurodevelopment of the children at 24-28 months was assessed with the Bayley Scales of Infant and Toddler Development, thirds edition (BSID-III) assessment (Bayley, 2006; Donald et al., 2018). The BSID-III is a well-validated tool that assesses child cognitive, language and motor development from 1-42 months and is sensitive to developmental delay (Bayley, 2006). The tool was administered by trained assessors with prompts in the child’s preferred language. Assessors were blinded to mother-child HIV status and were supervised throughout to ensure quality control. For the current study standardised composite scores were used. Composite scores were generated for each cognitive, language and motor domain, with a mean of 100 and standard deviation of 15, using normative United States data.

#### 2.2.3 Immune Assays

NGAL and MMP9 concentrations were measured using commercially available ELISA kits (NGAL: DY1757, MMP9: D911; R&D systems) in serum samples obtained from the mothers at 26 weeks gestation and the HEU children at 6-10 weeks and 24-28 months. Immune markers (GM-CSF, INF-γ, IL-1β, IL-2, IL-5, IL-6, IL-7, IL-8 and TNF-α, IL-4, IL-10, IL-12p70 and IL-13) were analyzed with a Milliplex® Luminex premix 13-plex kit (HSTCMAG28SPMX13; Merck) according to the manufacturer’s instructions. Plates were read on a Luminex system (Bio-Plex 200 System; Bio-Rad). All samples were assayed in duplicate.

### 2.3. Statistical Analysis

All data were tested for normality. Because of the skewed distribution, all markers were ln-transformed. This resulted in acceptable skewness and kurtosis of the data, which were used for further statistical analyses. Variables with missing values were: smoking status (n=8), alcohol use (n=14), socioeconomic status (n=5), BMI at 6 weeks (n=51), breastfeeding at 6-10 weeks (n=20), viral load (n=2), CD4+ count (n=19) and finally birth weight (n=2) and were considered missing at random. We therefore imputed values estimated from all predictor variables. Data were complete for all other covariates. Statistical analyses were performed on ten imputed datasets. The *Benjamini-Hochberg procedure* was used to control for the false discovery rate throughout the analyses due to multiple testing (McDonald, 2009).

First, unpaired t-tests were used to explore the differences of the inflammatory markers in mothers during pregnancy and children at 6-10 weeks and 24-28 months according to maternal HIV status. Significant correlations were subsequently used in linear regression analyses with the respective markers as dependent variables, maternal HIV status as predictor and adjusted for covariates. Pearson’s correlations were used to explore the associations between the inflammatory markers at each time point with neurodevelopment measures at 24-28 months of age. Multiple regression models were performed separately on significant findings with the respective inflammatory markers as dependent variables, neurodevelopment measure as predictor. Covariates were selected a priori based on their potential effects on either inflammatory markers or neurodevelopment. The following variables were added to the model: unadjusted; Model 1) maternal sociodemographic and lifestyle factors (adjusted for clinic, maternal smoking during pregnancy, maternal alcohol use during pregnancy, maternal socioeconomic status, maternal BMI at 6 weeks postpartum); model 2) infant health (adjusted for birth weight, prematurity, infant sex and exclusive breastfeeding (yes/no)); model 3) maternal HIV disease parameters (adjusted for maternal CD4+, maternal viral load during pregnancy and maternal ART regimen during pregnancy). All analyses were conducted using SPSS (version 25, IBM, USA). Group differences were considered statistically significant for all analyses where p-values were less than 0.05.

## 3. RESULTS

### 3.1. Participants

*Table 1* presents demographic data for mothers included in the study; sociodemographic characteristics were similar across comparison groups. There were significantly more HIV-infected women from Mbekweni compared to TC Newman clinic (*p<0.001*) as well as more HIV-infected mothers with moderate-severe alcohol exposure compared to HIV-uninfected mothers (*p=0.032*).

**Table 1:** Descriptive demographic characteristics for HIV-infected and HIV-uninfected mothers and neonatal measures at birth.

| **MOTHERS** | **HIV-infected** | **HIV-uninfected** | **P-value** |
| --- | --- | --- | --- |
| N (%) | 77 (28.8) | 190 (71.2) |  |
| Age, mean (SD) | 29 (5.43) | 26 (5.87) | 0.377 |
| Site, N (%) |  |  | **<0.001** |
| *Mbekweni* | 70 (90.9) | 70 (36.8) |  |
| *TC Newman* | 7 (9.1) | 120 (63.2) |  |
| Education, N (%) |  |  | 0.289 |
| *Primary* | 8 (10.4) | 14 (7.4) |  |
| *Some secondary* | 50 (64.9) | 108 (56.8) |  |
| *Secondary* | 17 (22.1) | 56 (29.5) |  |
| *Tertiary* | 2 (2.6) | 12 (6.3) |  |
| Married, N (%) |  |  | 0.933 |
| *Married/cohabitating* | 31 (39.2) | 76 (39.8) |  |
| *Other* | 48 (60.8) | 115 (60.2) |  |
| Body Mass Index (BMI), mean (SD) | 28.81 (6.60) | 26.23 (6.06) | 0.551 |
| Smoking (urine cotinine levels ng/ml), N (%) |  |  | 0.235 |
| *Non-smoker (<10 ng/ml)* | 21 (28.4) | 37 (20.0) |  |
| *Passive smoker (10-500 ng/ml)* | 31 (41.9) | 76 (41.1) |  |
| *Active smoker (>500 ng/ml)* | 22 (29.7) | 72 (38.5) |  |
| Alcohol exposure, N (%) |  |  | **0.032** |
| *Unexposed* | 61 (89.7) | 179 (96.8) |  |
| *Moderate-severe exposure* | 7 (10.3) | 6 (3.2) |  |
| Illness during pregnancy, N (%) | 11 (13.9) | 17 (8.9) | 0.213 |
| Maternal CD4+ count in pregnancy, mean (SD) | 453.50 (339.25-628.00) | - | - |
| Viral load during pregnancy, N (%) |  |  |  |
| *Below detectable limit (<40 copies/mL)* | 40 (51.9) | - | - |
| *Detectable (≥40-1000 copies/mL)* | 5 (6.5) | - | - |
| *Unsuppressed (>1000 copies/mL)* | 6 (7.8) | - | - |
| Antiretroviral regimen during pregnancy N (%) |  |  |  |
| *Prevention of mother-to-child transmission prophylaxis (zidovudine)* | 11 (14.3) | - | - |
| *First-line triple therapy* | 64 (83.1) | - | - |
| *Second-line or third-line therapy* | 2 (2.60) | - | - |
| Social-economic status (SES), N (%) |  |  | 0.475 |
| *Lowest SES* | 25 (32.5) | 43 (23.2) |  |
| *Low-moderate SES* | 17 (22.1) | 49 (26.5) |  |
| *Moderate-high SES* | 20 (26.0) | 55 (29.7) |  |
| *High SES* | 15 (19.5) | 38 (20.5) |  |
| Prematurity (<37 weeks), N (%) | 69 (89.6) | 164 (86.3) | 0.464 |
| Birth weight, mean (SD) | 3.02 (0.52) | 3.02 (0.59) | 0.202 |
| Delivery Method, N (%) |  |  | 0.052 |
| *Vaginal* | 48 (69.6) | 151 (83.4) |  |
| *Elective Cesarean* | 6 (8.7) | 9 (5.0) |  |
| *Emergency Cesarean* | 15 (21.7) | 21 (11.6) |  |

**SD: standard deviation, N: numbers.**

*Table 2* presents characteristics of HU and HEU children at 6-10 weeks and at 24-28 months. Both HEU and HU groups had similar growth indices at 6-10 weeks and 24-28 months. More HU infants were exclusively breastfed at 6-10 weeks compared to HEU infants (*p=0.032*). However, overall low rates of exclusive breastfeeding at 6-10 weeks occurred (45.5% in HEU and 60.8% in HU group), similar to findings across the cohort (Budree et al., 2017a). None of the children were exclusively breastfed at 24-28 months of age.

**Table 2:** Descriptive demographic characteristics for HEU and HU children at 6-10 weeks and 24-28 months.

| INFANTS 6 WEEKS | HEU | HUU | P-value |
| --- | --- | --- | --- |
| N (%) | 63 (71.6) | 159 (28.4) |  |
| Age (weeks), mean (SD) | 8.1 (1.5) | 7.9 (1.6) | 0.277 |
| Baby Sex (Female) N (%) | 37 (59.7) | 89 (57.1) | 0.723 |
| Weight, mean (SD) | 4.91 (0.70) | 4.83 (0.79) | 0.505 |
| Length, mean (SD) | 54.75 (2.66) | 54.67 (2.90) | 0.370 |
| Head circumference, mean (SD) | 38.86 (1.54) | 38.61 (1.81) | 0.197 |
| Exclusive breast feeding, N (%) | 30 (45.5) | 110 (60.8) | **0.032** |
| **CHILDREN 24 MONTHS** |  |  |  |
| N (%) | 77 (28.8) | 190 (71.2) |  |
| Age (months), mean (SD) | 27.3 (4.0) | 27.2 (3.5) | 0.331 |
| Baby Sex (Female) N (%) | 47 (61.0) | 113 (59.5) | 0.813 |
| Weight, mean (SD) | 11.98 (1.88) | 11.61 (1.63) | 0.249 |
| Length, mean (SD) | 83.40 (3.65) | 83.73 (3.54) | 0.889 |
| Head circumference, mean (SD) | 48.43 (2.15) | 47.96 (1.89) | 0.858 |

SD = Standard deviation, N = numbers

### 3.2. Inflammatory markers according to maternal HIV status

Serum levels of the inflammatory markers GM-CSF (*p=0.003) (Fig. 1A*) and MMP9 (*p<0.001) (Fig. 1B*) were significantly lower in HIV-infected compared to HIV-uninfected mothers at 26 weeks gestation. The inflammatory markers IL-1β (*p=0.021*) and IL-4 (*p=0.019*) were also lower in HIV-infected mothers (*p<0.05 uncorrected*), however this was not significant after multiple comparison correction.

**Figure 1:**
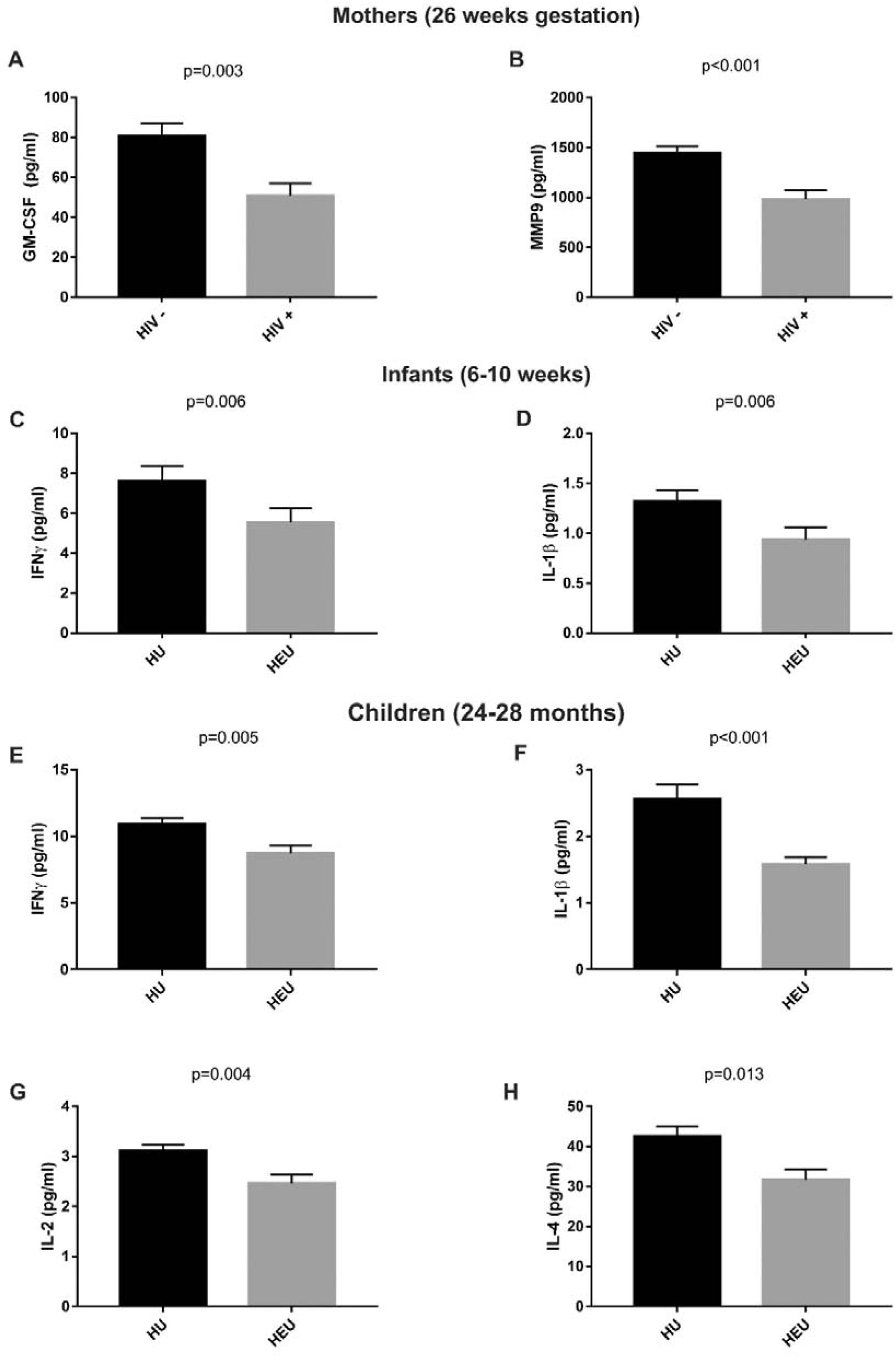
Comparisons of serum levels of all inflammatory markers that were significantly different in HIV-infected mothers and their HEU children compared to HIV uninfected mothers and HU children.

In infants at 6-10 weeks serum levels of the inflammatory markers IFN-γ (*p=0.006) (Fig. 1C*) and IL-1β (*p=0.006) (Fig. 1D*) were decreased in HEU compared to HU infants. The inflammatory markers IL-12p70 and IL-4 were lower in HEU infants (*p<0.05 uncorrected*) but this was not significant after multiple comparison correction.

In children at 24-28 months serum levels of inflammatory markers IFN-γ (*p=0.005) (Fig. 1E*), IL-1β (*p<0.001) (Fig. 1F*), IL-2 (*p=0.004) (Fig. 1G*) and IL-4 (*p=0.013) (Fig. 1H*) were all significantly lower in HEU compared with HU children after multiple comparison correction.

*Figure 1* indicates all inflammatory markers that were significantly reduced in HIV-infected mothers and their HEU children compared to HIV-uninfected mothers and their HU children. The natural log (ln) transformed values for all the inflammatory markers for the HIV-infected and HIV-uninfected mothers and their children are shown in the supplementary file (*supplementary Tables 1-3*).p=0.003

Multivariable linear regression analyses were performed for inflammatory markers according to maternal HIV status (*Table 3*). In adjusted analyses, GM-CSF (*p=0.016*) and MMP9 (*p=0.034*) was significantly lower in HIV-infected mothers. At 6-10 weeks, IFN-γ (*p=0.001*) was significantly lower in HEU compared to HU infants; IL-β (*p=0.120*) was not significant after controlling for all covariates. By 24-28 months, IFN-γ (*p=0.019*) IL-1β (*p=0.001*), IL-2 (*p=0.035*) and IL-4 (*p=0.017*) levels were significantly lower in HEU children after controlling for covariates.

**Table 3:** Multivariable linear regression analyses of inflammatory markers in mothers, infants (6-10 weeks) and children (24-28 months)

|  | B (SE) | B | P-value |
| --- | --- | --- | --- |
| **Mothers\*** |  |  |  |
| GM-CSF | -0.337 (0.14) | -0.171 | 0.016 |
| MMP-9 | -0.229 (0.11) | -0.144 | 0.034 |
| **Infants 6-10 weeks<sup>#</sup>** |  |  |  |
| IFN-γ | -0.566 (0.17) | -0.258 | 0.001 |
| IL-1β | -0.245 (0.16) | -0.120 | 0.120 |
| **Children 24-28 months<sup>†</sup>** |  |  |  |
| IFN-γ | -0.216 (0.10) | -0.168 | 0.019 |
| IL-1β | -0.401 (0.12) | -0.244 | 0.001 |
| IL-2 | -0.228 (0.11) | -0.150 | 0.035 |
| IL-4 | -0.366 (0.15) | -0.173 | 0.017 |

\***covariates:** clinic, smoking, alcohol exposure, age, socioeconomic status and body mass index (BMI) at 6 weeks postpartum.
<sup>#</sup>**covariates:** clinic, maternal smoking during pregnancy maternal alcohol exposure during pregnancy, maternal socioeconomic status and maternal BMI at 6 weeks postpartum, prematurity, birth weight, exclusive breastfeeding at 6-10 weeks and infant sex.
<sup>†</sup>**covariates:** clinic, maternal smoking during pregnancy, maternal alcohol exposure during pregnancy, prematurity, maternal socioeconomic status and maternal BMI at 6 weeks postpartum, birth weight and child sex.
**Abbreviations:** GM-CSF, Granulocyte-macrophage colony-stimulating factor; MMP-9 metalloproteinase-9; IFN-γ , interferon-γ ; IL, interleukin.

### 3.3. Association of inflammatory markers with neurodevelopment

No inflammatory markers in HIV-infected mothers were significantly associated with neurodevelopmental measures in HEU children after correction for multiple comparisons. However, markers of inflammation in HIV-infected mothers [IFN-γ (*r=-0.295, p=0.01*), IL-10 (*r=-0.253, p=0.036*), IL-12p70 (*r=-0.262, p=0.030*) and IL-7 (*r=-0.246, p=0.041*)] were associated with lower composite scores for language in HEU children (24-28 months) on initial analysis; but were not significant after multiple comparison correction. TNF-*α* (*r=-0.288, p=0.014*) was also associated initially with lower cognitive scores in HEU children (24-28 months) prior to multiple comparison correction (*Table 4*).

**Table 4:** Correlation between inflammatory markers in HIV-infected mothers and HEU children at 6-10 weeks and 24-28 months with neurodevelopment outcomes at 24-28 months.

| HIV-infected Mothers |  |  |  |  |  |  |  |  |  |  |  |  |  |  |  |  |
| --- | --- | --- | --- | --- | --- | --- | --- | --- | --- | --- | --- | --- | --- | --- | --- | --- |
| | | GM-CSF | IFN-γ | IL-10 | IL-12p70 | IL-13 | IL-1β | IL-2 | IL-4 | IL-5 | IL-6 | IL-7 | IL-8 | TNF-α | NGAL | MMP9 |
| Cognitive | *r* | .085 | -.131 | -.069 | -.045 | -.051 | -.023 | -.086 | .049 | -.128 | -.144 | -.226 | -.071 | **-.288** | -.016 | -.122 |
|  | P-value | .477 | .272 | .566 | .705 | .669 | .850 | .475 | .684 | .283 | .227 | .057 | .556 | **.014** | .892 | .307 |
| Language | *r* | -.142 | **-.295** | **-.253** | **-.262** | -.174 | -.223 | -.232 | -.208 | -.211 | -.204 | **-.246** | -.128 | -.206 | .081 | .045 |
|  | P-value | .245 | **.014** | **.036** | **.030** | .153 | .066 | .056 | .086 | .081 | .093 | **.041** | .295 | .089 | .509 | .711 |
| Motor | *r* | .031 | -.166 | -.147 | -.164 | -.082 | -.133 | -.085 | -.123 | -.134 | .012 | -.109 | -.029 | -.138 | -.036 | -.068 |
|  | P-value | .807 | .182 | .240 | .189 | .511 | .288 | .496 | .325 | .283 | .923 | .383 | .815 | .269 | .775 | .589 |
| HEU Infants (6-10 weeks) |  |  |  |  |  |  |  |  |  |  |  |  |  |  |  |  |
| | | GM-CSF | IFN-γ | IL-10 | IL-12p70 | IL-13 | IL-1β | IL-2 | IL-4 | IL-5 | IL-6 | IL-7 | IL-8 | TNF-α | NGAL | MMP9 |
| Cognitive | *r* | .005 | -.178 | -.136 | -.089 | -.091 | -.207 | -.126 | -.164 | -.010 | -.181 | -.238 | -.225 | -.178 | -.153 | -.186 |
|  | P-value | .971 | .178 | .305 | .501 | .492 | .116 | .343 | .214 | .938 | .171 | .070 | .086 | .177 | .251 | .162 |
| Language | *r* | .081 | -.106 | -.116 | -.055 | -.025 | -.199 | .007 | -.182 | -.070 | -.165 | -.061 | -.221 | -.152 | -.169 | **-.305** |
|  | P-value | .548 | .430 | .390 | .684 | .855 | .138 | .958 | .177 | .607 | .220 | .654 | .098 | .260 | .214 | **.022** |
| Motor | *r* | **-.309<sup>#</sup>** | **-.339<sup>#</sup>** | **-.451<sup>#</sup>** | **-.379<sup>#</sup>** | -.243 | **-.491<sup>#</sup>** | **-.308<sup>#</sup>** | **-.418<sup>#</sup>** | -.237 | **-.335<sup>#</sup>** | -.239 | -.003 | -.071 | **-.383<sup>#</sup>** | **-.289** |
|  | p-value | **.022** | **.011** | **.001** | **.004** | .074 | **.000** | **.022** | **.002** | .082 | **.012** | .079 | .981 | .607 | **.004** | **.034** |
| HEU Children (24-28 months) |  |  |  |  |  |  |  |  |  |  |  |  |  |  |  |  |
| | | GM-CSF | IFN-γ | IL-10 | IL-12p70 | IL-13 | IL-1β | IL-2 | IL-4 | IL-5 | IL-6 | IL-7 | IL-8 | TNF-α | NGAL | MMP9 |
| Cognitive | *r* | -.032 | -.191 | **-.303** | -.166 | .018 | -.148 | -.148 | -.118 | .013 | -.106 | -.177 | .010 | -.054 | .207 | -.007 |
|  | P-value | .793 | .111 | **.010** | .166 | .884 | .217 | .219 | .329 | .916 | .381 | .139 | .933 | .654 | .081 | .956 |
| Language | *r* | -.234 | **-.307** | **-.319** | **-.284** | .029 | **-.241** | **-.253** | -.217 | -.049 | -.012 | -.140 | -.025 | -.083 | .159 | -.001 |
|  | P-value | .055 | **.011** | **.008** | **.019** | .817 | **.048** | **.037** | .076 | .690 | .922 | .255 | .842 | .503 | .193 | .991 |
| Motor | *r* | -.050 | -.073 | -.215 | -.143 | .089 | -.124 | -.011 | -.154 | .100 | .091 | -.028 | .105 | -.027 | .112 | .023 |
|  | P-value | .690 | .562 | .086 | .256 | .480 | .326 | .934 | .221 | .429 | .473 | .825 | .405 | .830 | .371 | .856 |

In HEU infants (6-10 weeks) most inflammatory markers [GM-CSF (*r=10.309, p0.022*), IFN-γ (*r=-0.339, p=0.011*), IL-10 (*r=-0.451, p=0.001*), IL-12p70 (*r=-0.379, p=0.004*), IL-1β (*r=-0.491, p<0.000*), IL-2 (*r=-0.308, p=0.022*), IL-4 (*r=-0.418, p=0.002*), IL-6 and NGAL (*r=-0.383, p=0.004*)] were significantly associated with motor development in HEU children (24-28 months) after correction for multiple comparisons. MMP9 was shown to be initially associated with motor (*r=-0.289, p=0.034*) and language outcomes (*r=-0.305, p=0.022*) prior to multiple comparison correction. Additionally, IL-1β (*r=-0.342, p=0.008*) was associated with language outcomes prior to multiple comparison correction (*Table 4*).

No associations of inflammatory markers in HEU children (24-28 months) with neurodevelopment measures reached statistical significance after correction for multiple comparisons. However, in HEU children (24-28 months) IL-10 (*r=-0.303, p=0.010*) was associated with lower cognitive scores prior to multiple comparison correction. Inflammatory markers IFN-γ (*r=-0.307, p=0.011*), IL-10 (*r=-0.319, p=0.008*), IL-12p70 (*r=-0.284, p=0.019*), IL-1β (*r=-0.241, p=0.048*) and IL-2 (*r=-0.253, p=0.037*) were also associated with language outcomes prior to multiple comparison correction (*Table 4*).

There were no significant correlations between inflammatory markers in the HIV-uninfected mothers and their HU children with neurodevelopmental measures after multiple comparison corrections (*supplementary Tables 4-6*).

Multivariable linear regression analyses of inflammatory markers for infants at 6-10 weeks with motor development at 24-28 months showed that GM-CSF, IFN-γ, IL-10, IL-12p70, IL-1β, IL-2, IL-4, IL-6 and NGAL were associated with poorer motor development after controlling for covariates on all three models *(p<0.05) (Table 5)*.

**Table 5:** Multivariable linear regression comparing inflammatory markers for infants at 6-10 weeks and motor neurodevelopment scores at 24-28 months.

|  | Unadjusted |  |  | Model 1 |  |  | Model 2 |  |  | Model 3 |  |  |
| --- | --- | --- | --- | --- | --- | --- | --- | --- | --- | --- | --- | --- |
| | B (SE) | $\beta$ | P-value | B (SE) | $\beta$ | P-value | B (SE) | $\beta$ | P-value | B (SE) | $\beta$ | P-value |
| **GM-CSF** | -0.045 (0.02) | -0.309 | 0.022 | -0.048 (0.02) | -0.318 | 0.023 | -0.058 (0.02) | -0.395 | 0.003 | -0.058 (0.02) | -0.395 | 0.003 |
| **IFN-γ** | -0.035 (0.01) | -0.339 | 0.011 | -0.031 (0.02) | -0.285 | 0.047 | -0.037 (0.01) | -0.344 | 0.012 | -0.037 (0.01) | -0.345 | 0.012 |
| **IL-10** | -0.039 (0.01) | -0.451 | 0.001 | -0.039 (0.01) | -0.448 | 0.001 | -0.041 (0.01) | -0.470 | <0.001 | -0.041 (0.01) | -0.470 | <0.001 |
| **IL-12p70** | -0.038 (0.01) | -0.379 | 0.004 | -0.037 (0.01) | -0.359 | 0.010 | -0.041 (0.01) | -0.405 | 0.002 | -0.041 (0.01) | -0.405 | 0.002 |
| **IL-1β** | -0.042 (0.01) | -0.491 | <0.001 | -0.041 (0.01) | -0.488 | <0.001 | -0.043 (0.01) | -0.504 | <0.001 | -0.043 (0.01) | -0.504 | <0.001 |
| **IL-2** | -0.029 (0.01) | -0.308 | 0.022 | -0.031 (0.01) | -0.319 | 0.023 | -0.031 (0.01) | -0.320 | 0.021 | -0.031 (0.01) | -0.320 | 0.021 |
| **IL-4** | -0.050 (0.02) | -0.418 | 0.002 | -0.051 (0.02) | -0.406 | 0.003 | -0.053 (0.02) | -0.440 | 0.001 | -0.053 (0.02) | -0.440 | 0.001 |
| **IL-6** | -0.044 (0.02) | -0.335 | 0.012 | -0.052 (0.02) | -0.307 | 0.021 | -0.045 (0.02) | -0.337 | 0.012 | -0.045 (0.02) | -0.337 | 0.012 |
| **NGAL** | -0.020 (0.01) | -0.383 | 0.004 | -0.025 (0.01) | -0.451 | 0.001 | -0.021 (0.01) | -0.412 | 0.001 | -0.021 (0.01) | -0.412 | 0.001 |

**Model 1:** adjusted for clinic, maternal smoking during pregnancy, maternal alcohol use during pregnancy, maternal socioeconomic status, maternal body mass index (BMI) at 6 weeks postpartum.
**Model 2:** adjusted for birth weight, prematurity, exclusive breastfeeding at 6-10 weeks and infant sex.
**Model 3:** adjusted for maternal CD4+, maternal viral load during pregnancy and maternal antiretroviral therapy (ART) regime during pregnancy.
**Abbreviations:** GM-CSF, Granulocyte-macrophage colony-stimulating factor; IFN-γ , interferon-γ ; IL, interleukin; NGAL, neutrophil gelatinase associated lipocalin.

## 4. DISCUSSION

This is the first study reporting longitudinal associations between inflammatory markers in HIV-infected and uninfected pregnant mothers and their children with neurodevelopmental measures. Key findings were that maternal HIV infection was associated with decreased levels of inflammatory markers in pregnant women and in their children at 6-10 weeks and 24-28 months; there was a significant association between inflammatory markers in HEU infants at 6-10 weeks and poorer motor development at 24-28 months.

The results of this study are consistent with evidence indicating inflammatory cytokines that play an important role in immune regulation during pregnancy may be dysregulated in HIV-infected women. Reduced serum levels of inflammatory cytokines in HIV-infected mothers and HEU children were found, which were consistent with a few other studies. These studies found reduced levels of IL-2, IL-6 and TNF-α in HIV-infected mothers and reduced levels of IL-4, IL-7 and IL-12 in HEU infants (Borges-Almeida et al., 2011; Chougnet et al., 2000; Maharaj et al., 2017). However, the current study differs from most previously reported studies, which found increased levels of inflammatory markers (predominantly IL-1β, IL-6 and TNF-α) in both HIV-infected mothers and HEU children (Dirajlal-Fargo et al., 2019; Miyamoto et al., 2017; Richardson and Weinberg, 2011). The above studies were performed mostly in South America, America and European countries, which represent a population of women primarily infected with the HIV subtype Clade B; only two studies were in sub-Saharan Africa. The sub-Saharan epidemic of HIV infection, representing the greatest proportion of HIV-infected individuals and HEU children worldwide, is largely infected with the HIV subtype Clade C (Geretti, 2006). Clade C tends to present a more immunosuppressive profile, in keeping with our findings, compared to other clades such as Clade B, which have a pro-inflammatory effect with increased neuroinflammation possibly due to the amino-acid sequence differences of the Tat protein (Rao et al., 2013; Ruiz et al., 2019). Our finding of reduced serum levels of inflammatory markers associated with HIV infection in pregnant mothers and HEU children, highlights the potential dysregulatory impact of maternal HIV infection on immune function, particularly during the early critical period of child neurodevelopment. The impact of ART on levels of inflammatory markers in HIV-infected mothers and HEU children should also be considered, as this may reduce systemic inflammation and immune activation (Hileman and Funderburg, 2017). However, in our analysis, ART was included as a covariate, and the inflammatory environment of the HEU children was consistently low until 2 years of age when the children were no longer exposed to ART directly. Therefore, a significant impact of ART on cytokine levels in HEU children in this cohort appears less likely, although future research is needed to explore this area.

The reduction in cytokine serum levels in HEU children through the two years mirrors the reduced levels of cytokines found in their mothers, demonstrating a persistent effect. This suggests that the mothers’ immune profile may predict that of their children. Many studies have found that the maternal immune system significantly impacts the immune system of the foetus (Morelli S, 2015). One potential mechanism is through the transfer of maternal cytokines across the placenta (Abu-Raya et al., 2016b; Morelli S, 2015). There are very few studies that assess the inflammatory profile of HEU children longitudinally, highlighting the importance of this study and need for future investigation in this area.

Our results demonstrated that increased levels of several immune markers (GM-CSF, IFN-γ, IL-10, IL12p70, IL-1β, IL-2, IL-4, IL-6 and NGAL) in HEU infants at 6-10 weeks were associated with poorer motor development at 2 years. This suggests that an altered immune system early in life may predict neurodevelopmental delay in later childhood. A review by Bilbo *et al* supports this by highlighting the impact of the immune system on normal brain development (Bilbo and Schwarz, 2012). HEU infants may have a more sensitive immune system compared to HU infants and consequently this may result in increased susceptibility to changes in levels of inflammatory markers. HEU infants may manifest a lower threshold for the impact of fluctuations in inflammatory markers on the developing brain. This theory is supported by evidence that HIV exposure impacts the immune system of HEU children (Abu-Raya et al., 2016a). On the other hand, the immune systems of unexposed infants are assumed to be functioning under typical physiological conditions and therefore may be able to adapt more easily to fluctuations in inflammatory markers. Previous findings in animal models suggest that maternal immune compromise also affects neurodevelopment via changes in foetal inflammatory markers (Yockey and Iwasaki, 2018).

Poorer language functioning in HEU children was associated with increased serum levels of IFN-γ, IL-10 and IL-12p70 in HIV-infected mothers and their children before multiple comparison correction. This finding, though not reaching statistical significance remains important to note, as studies reporting on associations between HEU and early development have consistently found that early language outcomes are affected (Le Doare et al., 2012). We recently reported that HEU children in the DCHS (Wedderburn et al., 2019) showed significantly lower language scores compared with HU children at 2 years of age. Further investigation into the impact of HIV exposure and mechanisms for impact on language neurodevelopment is needed.

Limitations of this study include the modest sample size warranting replication in a larger study; however, this is one of the largest studies to investigate inflammatory markers longitudinally in HEU and HU children. Although the study population is characteristic of this region of South Africa and many other parts of sub-Saharan Africa, care must be taken when extrapolating the results to other environments. The findings may not apply to populations with HIV clades other than HIV clade-C, which is the predominant clade in sub-Saharan Africa, as HIV subtypes may affect immune markers (Gandhi et al., 2009).

## 5. CONCLUSION

This follow-up study demonstrates that maternal HIV infection is associated with immune dysregulation; with results indicating suppressed levels of serum inflammatory markers in HIV infected mothers and HEU children through to two years of age. The results further show that an altered immune system in HEU infants is associated with poorer motor development in children at two years. This study adds to the emerging understanding of the role of HIV and the immune system in neurodevelopment. Further, the findings suggest potential mechanisms through which early interventions may be developed in order to support optimal neurodevelopmental outcomes in HEU children across their lifespan.

## Data Availability

On request

## 6. ACKNOWLEDGEMENTS

The Drakenstein Child Health Study was funded by the Bill & Melinda Gates Foundation (OPP 1017641), Discovery Foundation, Medical Research Council South Africa, National Research Foundation South Africa, CIDRI Clinical Fellowship and Wellcome Trust (204755/2/16/z). DJS, HJZ, KAD and WB are supported by the SA Medical Research Council (SAMRC). CJW is supported by the Wellcome Trust through a Research Training Fellowship [203525/Z/16/Z]. WB is supported by the SAMRC, through its Division of Research Capacity Development under the Bongani Mayosi National Health Scholars programme. PJWN is supported by the National Alliance for Research on Schizophrenia and Depression Young Investigator Grant (No. 25199) and Scott-Gentle Foundation. The funders had no role in the study design, data collection and analysis, decision to publish, or preparation of manuscript.

